# Quantitative longitudinal natural history of eight gangliosidoses – conceptual framework and baseline data of the German 8-in-1 disease registry. A cross-sectional analysis

**DOI:** 10.1101/2022.04.13.22273562

**Authors:** Markus Ries, Grecia Mendoza, Laila Arash-Kaps, Yasmina Amraoui, Folker Quack, Brigitte Hardt, Stefan Diederich, Michael Beck, Eugen Mengel

## Abstract

**Purpose:** Gangliosidoses are a group of inherited neurogenetic autosomal recessive lysosomal storage disorders usually presenting with progressive macrocephaly, developmental delay and regression, leading to significant morbidity, and premature death. A quantitative definition of the natural history would support and enable clinical development of specific therapies.

**Methods:** Single disease registry of eight gangliosidoses (NCT04624789).

Cross-sectional analysis of baseline data in N= 26 patients.

Primary endpoint: disease severity assessed by the 8-in-1 score.

Secondary endpoints: first neurological sign or symptom observed a. by parents and b. by physicians, diagnostic delay, as well as phenotypical characterization.

Tertiary endpoints: Neurological outcomes (development, ataxia, dexterity) and disability.

**Results:** The 8-in-1 score quantitatively captured severity of disease. Parents recognized initial manifestations (startle reactions) earlier than physicians (motor developmental delay and hypotonia). Median diagnostic delay was 3.16 [IQR 0.69 … 6.25] years. Eight patients presented with late-infantile phenotypes.

**Conclusion:** Data in this registry raise awareness of these rare and fatal conditions in order to accelerate diagnosis, inform counselling of afflicted families, define quantitative endpoints for clinical trials, and can serve as historical controls for future therapeutic studies. The characterization of a late-infantile phenotype is novel. Longitudinal follow-up is planned.

## Introduction

Gangliosidoses are rare, autosomal recessively inherited lysosomal storage disorders. The inability to degrade complex glykolipids due to the deficiency of certain enzymes and other proteins in the lysosome leads to the accumulation of gangliosides in various tissues. There are eight different gangliosidoses, defined by biochemical and genetic features. They can be grouped into two groups of four diseases respectively, i.e. the G_M1_ gangliosidosis / sialidosis group (N=4 diseases) and the G_M2_ gangliosidosis group (N=4 diseases), Supplemental Table 1. ^1-6^

These conditions usually present as global developmental delay and regression associated with distinct features such as macrocephaly, cherry-red spots, muscular hypotonia, ataxia, as well as exaggerated startle responses to sounds, light flashes and tactile stimuli. ^1-5^ Patients with attenuated phenotypes may not show all classical manifestations which lowers the degree of the clinician’s clinical suspicion. These individuals may be more difficult to diagnose which may mean that the time to diagnosis is delayed. It is also possible that some adult patients are never identified.^7,8^

Due to the rare nature of these conditions, the precise natural history of gangliosidoses is incompletely known. A quantitative understanding will inform the development of therapeutic interventions with regard to endpoints and the degree of their variability. ^7^ A precise knowledge of the natural history is helpful for counselling afflicted families and planning future therapeutic clinical trials. Therefore, research on the natural history of rare diseases was defined as a crucial element to advance therapeutic developments by the International Rare Diseases Research Consortium. ^9^ An independent governance of a disease registry for pre-and post-approval of novel treatments for rare diseases is important for the community of healthcare professionals, patients, regulators as well as for the pharmaceutical industry. ^10^ The purpose of this 8-in-1 independent German gangliosidoses registry is to quantitatively document the natural history of eight gangliosidoses in order to raise awareness of the diseases and to better understand these conditions.

We therefore directed our efforts in developing a comprehensive, independent national gangliosidoses disease registry in Germany jointly funded by the patients’ advocacy organization “Hand in Hand e.V.” and the independent non-profit clinical research organization “SphinCS Lyso gemeinnützige UG”. We chose an inclusive and participatory approach. The patients’ advocacy organization was directly involved in design and feasibility assessments of the registry in order to ensure that the patients’ and families’ perspectives are taken into account and represented in this project.

The conceptual framework, operationalization, and a baseline analysis of the 8-in-1 gangliosidoses registry’s natural history data will be presented here. We will primarily focus on quantification of disease severity as this item is crucial for understanding the outcome of the disease and will be of longitudinal interest in the future. Patients and afflicted families’ perspectives are a very important element of this project. Their perspectives on the disease will be juxtaposed to the healthcare professionals’ views in terms of first neurological manifestation. Diagnostic delay may be an issue and will be assessed for the present cohort (Table 1).

**Table 1:** Synopsis of quantitative natural history study assessments in the 8-in-1 disease registry

| Focus of assessment | Instrument | Study endpoint | Hypotheses |
| --- | --- | --- | --- |
| Disease severity | 8-in-1 score | Primary | 1) Older patients with the same subtype have higher disease burden scores because the conditions are progressive over time |
| First neurological sign or symptom observed by parents and physician | Medical history for first neurological sign or symptom | Secondary | 2) Health care professionals and parents assess time and seminal pattern of disease differently because of independent perspectives and assessment settings |
| Diagnostic delay | Time between onset of disease and time of diagnosis | Secondary | 3) There is a substantial diagnostic delay because gangliosidoses are rare diseases |
| Phenotypical characterization | General medical history | Secondary |  |
| Phenotypical characterization | Physical examination | Secondary |  |
| Phenotypical characterization | Neurological examination | Secondary |  |
| Phenotypical characterization | Subtype classification | Secondary |  |
| Development | Vineland adaptive behavior scales | Secondary |  |
| **Ataxia** | SARA score | Tertiary |  |
| **Dexterity** | Nine Hole Peg Test (NHPT) | Tertiary |  |
| **Disability** | WHO disability assessment schedule 2.0 | Tertiary |  |
| **Biomarker** | Clinical routine laboratory (CBC, clinical chemistry) and biobanking | Tertiary |  |

## Material and Methods

STROBE criteria were respected. ^11^ This study was approved by the state medical board Hessen, Frankfurt, Germany (registration number 2019-1483-evBO) and registered on clinicaltrials.gov (NCT04624789). All parents, caregivers, or patients signed informed consent or, if applicable, assent before inclusion into the study.

Inclusion criteria were 1) biochemically and/or genetically confirmed diagnosis of a gangliosidosis and 2) written informed consent. Exclusion criteria were 1) the absence of a biochemically and/or genetically confirmed diagnosis of a gangliosidosis or 2) the inability or unwillingness to give written informed consent. Pertinent European and German data protection laws and regulations were respected. An overview of the study assessments, instruments, and endpoints is provided in Table 1.

### Primary endpoint

Due to the absence of a disease specific instrument which captures disease severity and disease progression of patients with gangliosidoses, we developed the 8-in-1 composite impairment score adapted from similar scores in clinically related neurodegenerative and lysosomal storage diseases, i.e., NPC and CLN. ^12,13^ The 8-in-1 score summarizes eight domains of impairment, i.e., 1) participation, 2) care/ independency, 3) ambulation, 4) dexterity/ fine motor skills, 5) speech, 6) swallowing, 7) epilepsy and 8) cognition. Each domain can be scored from 0 to 4. Definitions and operational guidance are provided in Supplemental Table 2. The summation of all scores of each domain yields the total score, ranging from 0 to 32, with a higher score indicating more severe clinical impairment. As this is a cross-sectional analysis of the baseline data, no prospective data about disease progression is available yet and we defined the disease severity assessed by the 8-in-1 composite impairment score as primary endpoint. For each patient the 8-in-1 score was completed by the investigator and the caregiver independently.

### Secondary endpoints

Secondary endpoints include onset and nature of first neurological sign or symptom. Observations of parents and physicians were captured separately and compared, because parents’ and physicians’ ascertainments and perceptions can vary.

### Comparison time of onset parents vs. physicians

We hypothesize that patients recognize pertinent features earlier than physicians, although they do not perceive this as a symptom of a disease. It is possible that there is an early pattern of unspecific manifestations that parents are more susceptible to. Patients were classified into disease subtypes.^14,15^ In addition, we considered the disease onset above 6 months and below 2 years in G_M2_ as late-infantile, because this age span is not specifically addressed in Toro et al. ^15^

Regarding their phenotypes, two patients (patients 12 and 17) could not be categorized according current phenotype classification.^15^ These two patients had their first symptom before age of 6 months, but they reached higher “best performance” than seen for patients with late-infantile phenotype. Therefore, we classified them as late-infantile phenotype.

Medical history was captured through interviews and available reports in all patients. A physical and neurological examination was done in all patients except patients 2, 6, and 7 who participated in another clinical trial and patient 4 (assessed by videoconference).

Terminology for motor development was used as proposed by Piper and Darrah. ^16^

Development was assessed by the Vineland Adaptive Behavior Scales (VABS) (Vineland-3). This is a standardized assessment tool that utilizes semi-structured interviews to measure adaptive behavior and support the diagnosis of conditions such as intellectual and developmental disabilities, autism, and developmental delays. ^17^ The age range of the instrument covers birth to 90 years. Parents and caregiver interviews were considered, because this instrument is not suitable for patient interviews in individuals with severe cognitive impairment.

In order to systematically categorize and illustrate developmental delay and regression in gross motor development and based on our extensive clinical experience, the following six phases were defined:

#### First phase

free of motor signs and symptoms (time from birth until first gross motor abnormality).

#### Second phase

delay in gross motor function (time from first gross motor abnormality to initial loss of gross motor function).

#### Third phase

initial phase of gross motor regression, e.g., development of clumsy and/or atactic gait, but free walking (time from first loss of gross motor function until walking with assistance).

#### Fourth phase

Assisted walking (time from walking with assistance until complete loss of walking with assistance resulting in being wheel-chair bound).

#### Fifth phase

No autonomous locomotion (patient is wheelchair bound, but can still lift arms and legs from a surface).

#### Sixth phase

Minimal or no gross motor abilities (arms and legs cannot be lifted from surface anymore).

### Tertiary, i.e. exploratory endpoints

Because ataxia can cause clinically relevant impairment in patients with gangliosidosis, the neurological disease progression was assessed by the **Scale for the Assessment and Rating of Ataxia (SARA)**. SARA is a composit, 8-item rating scale based on a semiquantitative assessment of cerebellar ataxia on an impairment level and assesses clinical functions pertinent to ataxia, i.e., 1) gait, 2) stance, 3) sitting, 4) speech, 5) finger chase, 6) nose-finger test, 7) fast alternating hand movements, and 8) heel-shin slide. The numeric summary scale ranges from 0 to 40, with “0” meaning no ataxia and “40” being the worst ataxia. SARA was originally developed for the assessment of ataxia in adults and validated in two studies of N=167 and N=119 patients with spinocerebellar ataxia. ^18,19^ In addition, this instrument was used to assess the effects of acetyl-dl-leucin on ataxia in adolescent and young adult patients with Niemann-Pick type C. ^20^ In the present paper, SARA was used for patients, who were older than 8 years of age and who were able to follow instructions.

The bimanual dexterity was assessed by the Nine-Hole Peg Test (NHPT, AFH Webshop, Germany). The NHPT was originally introduced as a measure of dexterity ^21^, detailed test instructions and adult normative values according to hand, sex, and age are available. ^22^ Patients aged ≥ 5 years were asked to pick up the pegs from a container one at a time, place the pegs into the holes in any order until all the holes were filled, and then remove the pegs one at a time and return them to the container, all as quickly as possible. The time started as soon as the subject touched the first peg and stopped when the last peg hit the container. Both the dominant and nondominant hands were tested once. The time delay (in seconds) in dexterity performance of the dominant hand compared to age- and sex- referenced means of normal was determined. ^23^

In order to quantitate the burden of disability, the six functional domains 1) cognition, 2) mobility, 3) self-care, 4) getting along, 5) life activity, and 6) participation were assessed by the WHO disability assessment (WHODAS) 2.0 36 item version in adult patients only, because this instrument is validated for the adult population. ^24^ WHODAS 2.0 is a generic scoring instrument for health and disability used across cultures and languages in variety of diseases including neurological conditions. ^25^ To each of the six functional domains, a 5-level ordinal item-score ranging from “none” (0) to “extreme” (4) is attributed. The sum of all item-scores ranging from 0 to 144 describes the degree of functional limitation in an individual patient, with a higher score indicating more severe functional impairment.

The protocol allows the collection of blood for future analysis of potential biomarkers.

### Statistics

Standard techniques of descriptive statistics were applied. Categorical variables were summarized with frequencies and percentages. Continuous variables were summarized with mean, standard deviation, median, minimum and maximum values. Correlations between variables were assessed by Pearsons’ r. Missing data were not imputed.

The 8-in-1 score was cross-sectionally assessed against age and disease subtype. The goal was to analyze whether disease burden, as assessed by the 8-in-1 score in this progressive disease, was higher in older patients compared with younger patients. In addition, we analyzed whether the cross-sectional pattern of disease progression suggested either a slower or a faster course depending on the disease subtype, i.e., infantile onset patients should progress faster than adult-onset patients according to our observations in clinical practice.

Inter-group rater variability of 8-in-1 scored were compared between health care professionals and caregivers, because both groups may have a different perspective on the individual patients. 8-in-1 scores attributed by healthcare professionals and caregivers were compared with a Wilcoxon matched-pairs signed rank test in a conservative approach as the data did not have a Gaussian distribution. P values below 0.05 were considered statistically significant.

Ages at motor developmental delay and motor regression in patients with gangliosides stratified by disease and subtype were visualized graphically and calculated by summary statistics. We focused on motor function because this information was ascertained retrospectively and would therefore be less prone to recall bias than, for example the items speech and cognitive development which parents remember less well. Diagnostic delay was calculated as the difference between age of first neurological sign or symptom noted by parents and age at diagnosis.

We used GraphPad Prism 9 for Windows 64–bit, Version 9.1.0 (March 15, 2021, GraphPad Software, LLC, www.graphpad.com) to perform statistical calculations and to draw graphs.

## Results

### Patient population

In this analysis, we included 26 patients overall, referred to our institution as a convenience sample. Four patients had G_M1_ - gangliosidosis, and 22 patients had G_M2_ - gangliosidosis. Of the 22 patients with G_M2_ - gangliosidosis, 13 patients had Tay-Sachs disease, 7 had Sandhoff disease, and 2 had a GM2 activator deficiency (Supplemental Table 3, available upon request).

Three patients were included solely in the retrospective part of this study, because they were participating in a clinical trial with an investigational drug before the enrolment in our registry which would confound their natural history assessments.

### Primary endpoint: 8-in-1 score

Cross-sectional assessment of disease severity by 8-in-1 scores against age

Younger patients had lower 8-in-1 scores than older patients (Figure 1). When clustered by disease subtype (I.e., infantile, late infantile, juvenile, or adult onset across disease entities), patients with late-onset subtypes had relatively lower 8-in-1 scores at a later age compared with patients with more acute disease onset who reached higher 8-in-1 scores at a relatively younger age.

**Figure 1:**
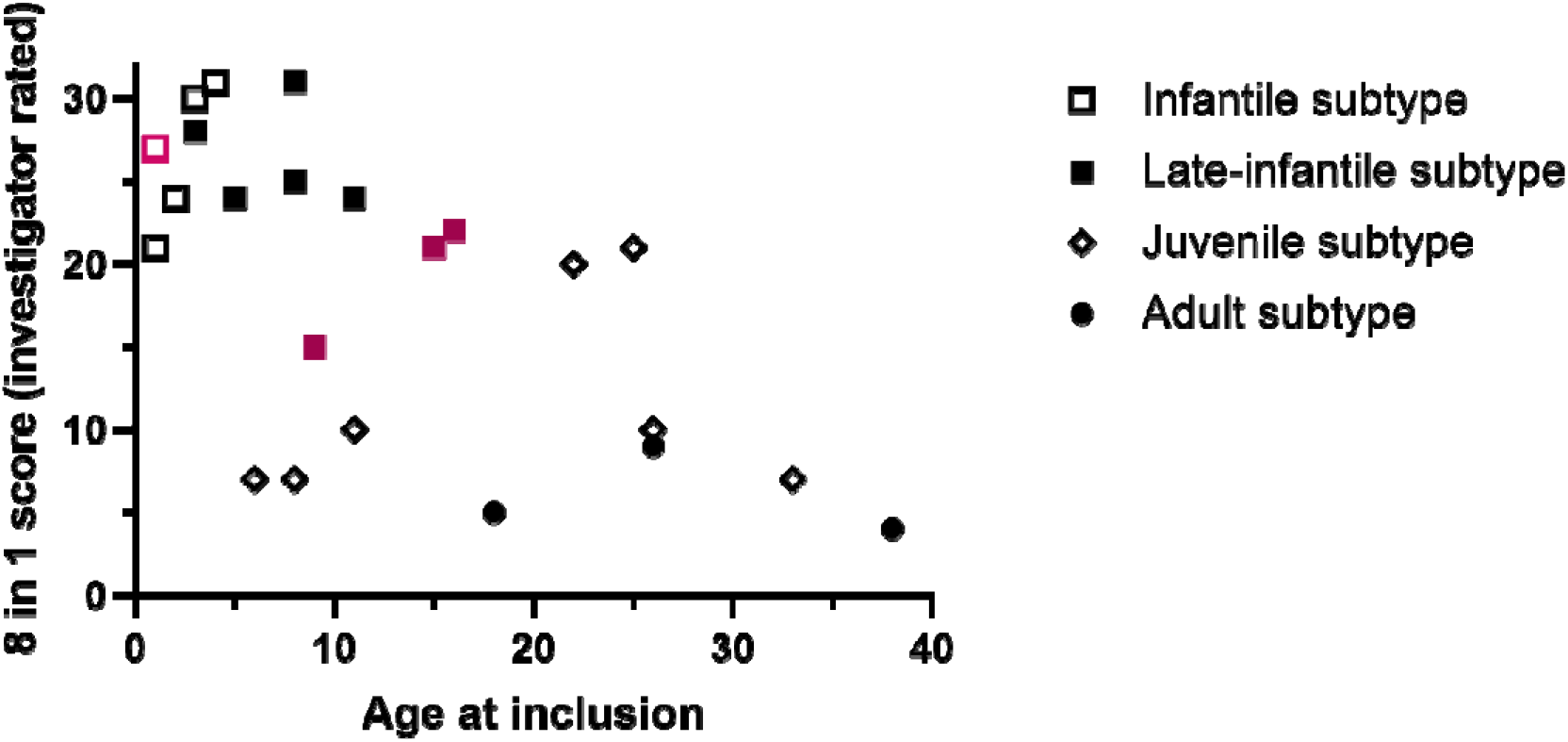
Disease severity cross-sectionally assessed by 8-in-1 score as a function of age and disease subtype gangliosidosis. Black indicates patients with G_M2_ – gangliosidosis, red indicates patients with G_M1_ – gangliosidosis.

Disease severity inter-group rater variability healthcare professionals vs. caregivers

Disease severity assessed with the 8-in-1 score in a given patient was rated higher by health care professional compared with ratings by caregivers (p=0.0383), Wilcoxon matched pairs-signed rank test, N=23), Figure 2.)

**Figure 2:**
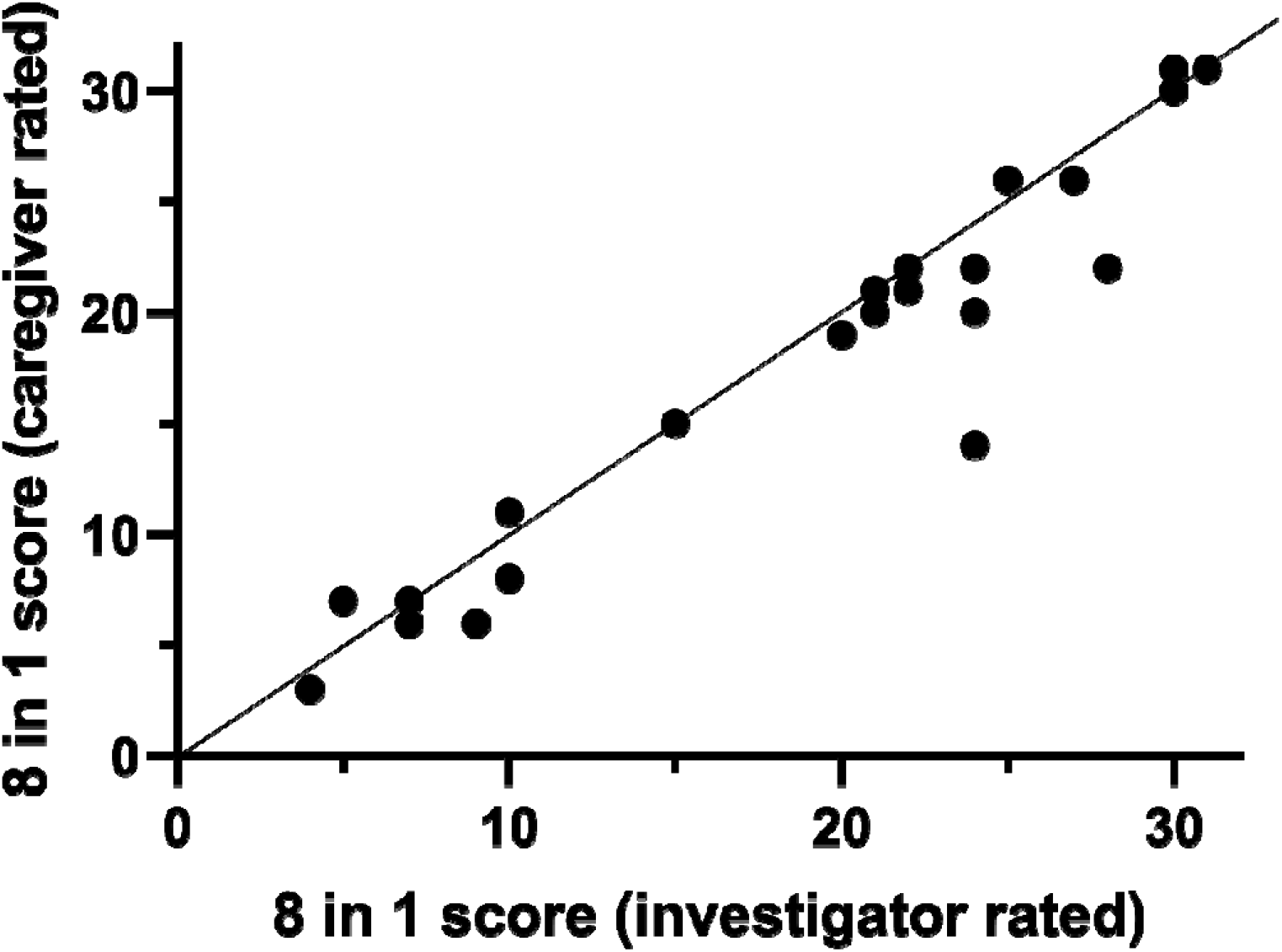
Inter-group rater variability comparing disease severity assessed by 8-in-1 scores attributed by 1) healthcare professionals and 2) caregivers Disease severity inter-group rater variability healthcare professionals vs. caregivers Disease severity assessed with the 8-in-1 score in a given patient was rated higher by health care professional compared with ratings by caregivers (p=0.0383, Wilcoxon matched-pairs signed rank test, N=23), figure.

Comparison of 8-in-1 scores with previously validated instruments of cognitive development (secondary endpoint), ataxia, dexterity, and disability (tertiary endpoint).

8-in-1 scores showed a high degree of correlation with *Vineland score for cognition* (Pearsons’ r = -0.5312; p=0.0110), and *SARA-scores* for motors function and coordination (Pearsons’ r = 0.6806; p=0.0117) whereas correlations with 9HPT (Pearsons’ r = 0.4293; p=NS) and WHODAS (Pearsons’ r = 0.6929; p=NS) were not statistically significant (Supplemental Figure 1).

### Secondary

First neurological sign or symptom observed by parents and physicians

Parents recognized abnormalities earlier than physicians. Median age at first sign/symptom recognized by parents was 1.850 [IQR 0.24…4.25] months for the overall group vs. 3.3 [IQR 1.05…6.5] months of age as detected by physicians (p<0.0001, Wilcoxon signed rank test).

In the infantile and late infantile types, parents recognized mainly startle reactions and poor fixation as first manifestation (Supplemental Table 3, available upon request). In juvenile patients, parents did also see startle reactions, but described mainly a predominantly heterogenous spectrum of impaired gross motor function as first manifestations.

In contrast, physicians mainly saw motor developmental delay and/or hypotonia as first manifestation in infantile patients. In juvenile patients, physicians reported ataxia and weakness in the lower extremities as the predominant first sign. In adult patients, parents’ and physicians’ observations were similar and included attention deficit and impaired function of the lower extremities.

### Diagnostic delay

Median diagnostic delay, i.e., the time between observation of first neurological sign or symptom seen by parents and the final diagnosis, was 3.16 [IQR 0.96 6.25] years for the overall group. Likewise, the diagnostic delays in the infantile, late-infantile, juvenile, and adult groups were 0.92 [IQR 0.5 … 1.165], 2.54 [IQR 0.46 … 3.475], 5.835 [IQR 3.375 … 19], and 5.5 [IQR 4.25 16.5] years (Figure 3)

**Figure 3.**
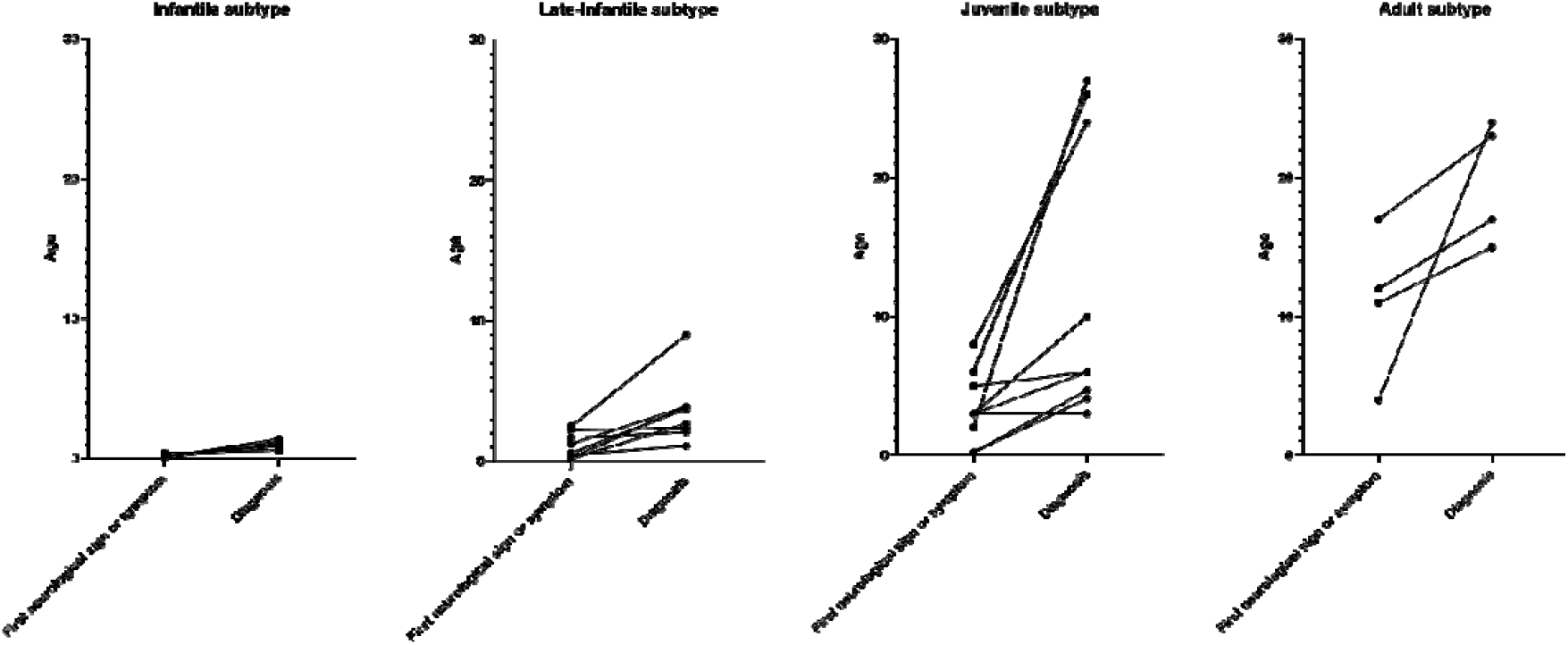
Diagnostic delay by disease subtype, i.e., the time between observation of first neurological sign or symptom seen by parents and the final diagnosis.

### Phenotypical characterization

Onset of disease and progression in individual patients is illustrated in Figure 4.

**Figure 4:**
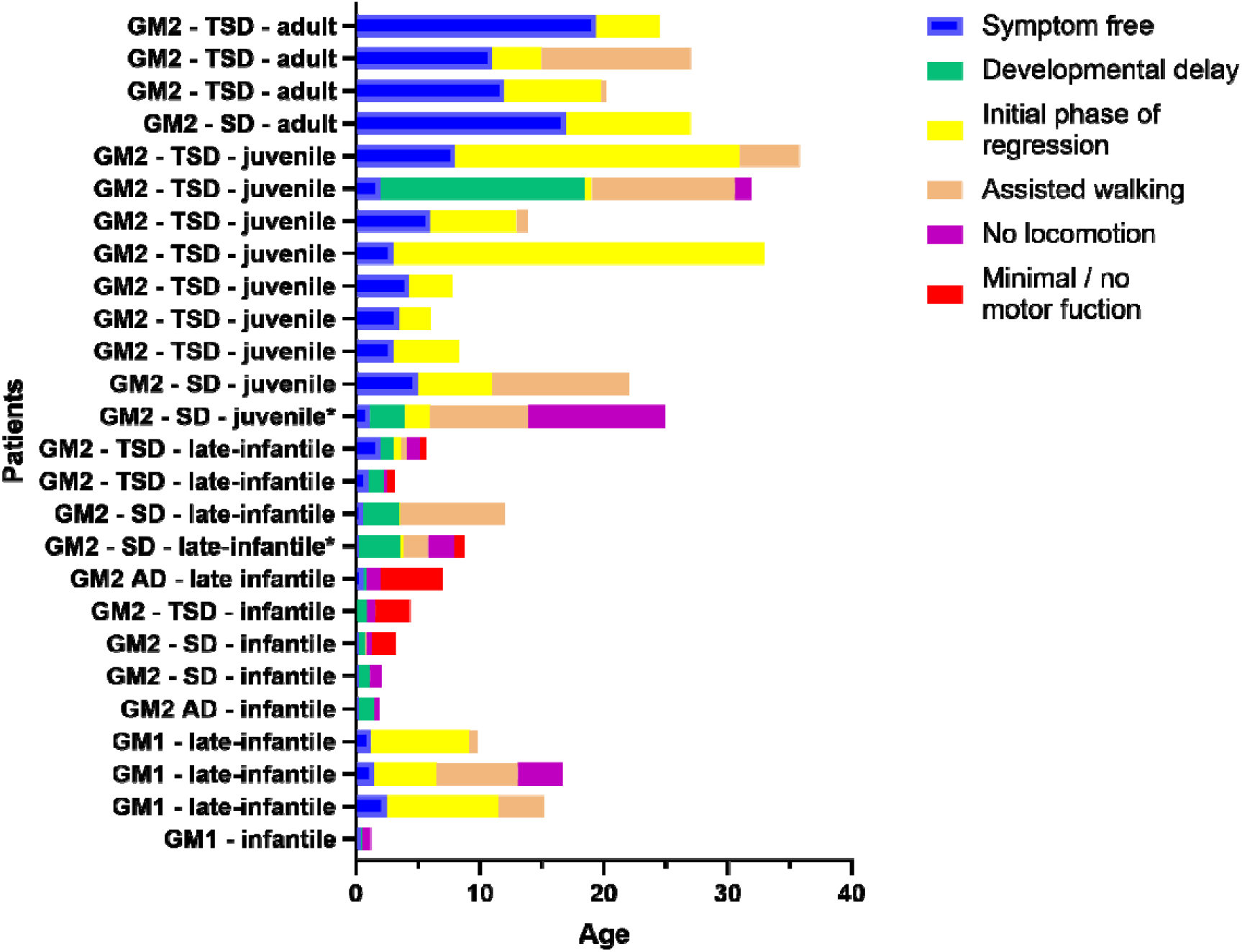
Retrospective assessment of ages at motor developmental delay and motor regression in patients with gangliosides, stratified by disease and subtype.

In qualitative terms, infantile patients are free of signs and symptoms for a short period only, followed by a relatively rapid progression into severe disease compared with the other less acute or late-onset subtypes. In contrast, adult-onset patients are free of signs and symptoms for a considerable period of time and their disease progression is then relatively mild and slow compared with the early onset subtypes. Late infantile patients with G_M2_ - gangliosidoses (N=5) tended to have a more acute onset and rapid decline compared with late infantile patients with G_M1_ - gangliosidosis (N=3)

### Development

Supplemental Table 3 (available upon request) shows data on development as assessed by Vineland scores. All patients had a substantial development disorder. Motor skills were compromised most significantly, whereas patients showed a relative strength in the socialization domain.

### Tertiary endpoints

#### Ataxia

SARA scores were determined in 10/26 patients (Supplemental Table 3, available upon request). The items gait, speech, and heel-chin slide appear to be affected more predominantly as assessed by SARA scores, compared with the other three remaining items, that were relatively preserved.

#### Dexterity

Seven out of 26 patients were eligible or able to complete the 9-Hole Peg Test, the results are summarized in Supplemental Table 3 (available upon request). All showed dexterity performance below the mean reference ranges.

#### Disability

Quantitation of disability by WHODAS 2.0 was feasible in a small subset of adult patients (N=6/26 patients overall). Four patients showed a moderate to severe degree of disability whereas two patients were less severely affected (Supplemental Table 3, available upon request).

## Sequence variants

Twenty-seven various gene variants of the *HEXA-, HEXB-GM2A-* and *GLB1-gene* were reported in 21 patients with 26 of them previously described in the literature. In one patient with Sandhoff disease (patient 20), a previously unreported homozygous variant was detected in exon 1 of the *HEXB* gene: c.149-158del; p.Ala 50Glyfs*11. This frameshift is causing a premature stop-codon and was classified likely pathogenic. Mutation taster predicts a disease-causing amino acid exchange. ExAC-Browser, Varsome and AdGenom did not list this sequence variant.

## Discussion

We present a comprehensive quantitative description of the broad spectrum of clinical manifestations in a large cohort of 26 patients with G_M1_- and G_M2_-gangliosidoses, covering the time from onset of disease, the medical history up to a thorough cross-sectional clinical characterization as baseline for future longitudinal natural history follow-up studies. This includes investigations with novel disease specific assessment tools such as the 8-in-1 disease score which quantitatively documents the severity of the disease in an individual patient.

The 8-in-1 score captured the different cross-sectional disease evolution patterns in the respective subgroups. The cross-sectional pattern of 8-in-1 scores as a function of age suggested that disease burden in gangliosidoses increased over time. In addition, the disease subtype appeared to predict progression, i.e., patients with infantile subtypes were relatively sicker compared with late-onset patients at a given age. Disease severity was highest in the early onset phenotypes, and, within a given phenotype, higher in the older patients. Therefore, the 8-in-1 score appeared to capture increasing disease severity as a function of age in these progressive conditions. We observed interesting differences between G_M1_ - and G_M2_ – isosubtypes: late infantile GM2 patients (N=4) had more severe disease than late infantile G_M1_ patients (N=3) (Figure 1). Furthermore, late infantile G_M2_ patients tended to progress into developmental regression faster than late infantile patients with G_M1_-gangliosidosis (Figure 4). However, this preliminary finding is limited by the small number of late infantile patients (N=8). There were no juvenile and adult patients with G_M1_-gangliosidoses for comparison with our subgroups of patients with juvenile and adult-onset patients with G_M2_-gangliosidoses.

In general, health care professionals rated disease severity slightly higher than caregivers (Figure 2), in particular, parents did not recognize seizures in two patients which accounted for the difference. 8-in-1 scores showed a statistically significant correlation with SARA scores (assessing ataxia) and Vineland adaptive behavior scales (assessing development). SARA scores correlated best with the 8- in-1 score (Pearson’s r = 0.8250), the correlation with the Vineland scores was -0.5312.

Testing with 9HPT was too difficult for the majority of patients, because they were too young or too severely affected and could therefore not perform the assessments. Results showed a high degree of variability and did not correlate with overall disease assessed by the 8-in-1 score. We therefore do not recommend using this tool in clinical studies with gangliosidoses.

The WHODAS 2.0 instrument was able to quantitate disability in a subset of patients. The instrument was not feasible for the majority of patients because it is validated in the adult population only. The general distribution pattern of these scores was similar to other neurodegenerative disorders, such as multiple sclerosis.^26^ The baseline scores documented in this publication will be followed up longitudinally in order to describe the further natural history course of disability in gangliosides in those patients for whom WHODAS 2.0 assessment can be performed.

The adult SARA score contains eight items to assess ataxia. In G_M2_-gangliosidoses, the items gait and heel-shin slide are particularly compromised due to distal limb paresis of the lower extremities as part of the underlying condition. In contrast, according to our clinical experience, function of the upper extremities such as hand and finger coordination usually function better and are longer preserved during the early course of the disease.

The perception of first signs and/or symptoms differed between parents and physicians. In the infantile and late infantile types, parents recognized mainly startle reactions and poor fixation as first manifestation, whereas physicians mainly saw motor developmental delay and/or hypotonia as first manifestation. In juvenile patients, parents also noticed startle reactions, but mainly reported a heterogenous spectrum of impaired gross motor function as first manifestations. Physicians noticed ataxia and weakness in the lower extremities as the predominant first sign in juvenile patients. In adult patients, parents’ and physicians’ observations were similar and included attention deficit and impaired function of the lower extremities. This could mean that cerebellar impairment is less important within the initial disease manifestations in adult patients compared with juvenile patients. Startle reactions, in contrast, were a typical first manifestations recognized by parents, but not physicians, in the infantile and late infantile group, and, to a lesser degree, in juvenile patients. Diagnostic delay was substantial which may be due to the fact that gangliosidoses are rare disorders. Targeted diagnostics are usually initiated only when the loss of abilities suggests a neurodegenerative disease. Increased disease awareness among health care providers or easier access to powerful diagnostic tools such as whole exome sequencing may improve the time to diagnosis.

There are natural history studies and reviews in the literature that document onset of disease and neurological manifestations in gangliosidoses. These valuable reports focus mainly on a specific clinical, genetic, or imaging aspect or a distinct subtype of gangliosidoses, but none of these studies encompass the holistic overall spectrum of gangliosidoses in a quantitative way. The present data help to close this important gap. Of interest, two studies analyzed patients with G_M1_ - and G_M2_ - gangliosidoses in a comparative way which corroborates the methodology of the present registry.^27,28^

### Limitations and directions for future research

The present study has some limitations. First, the present population is skewed towards G_M2_-gangliosidoses, which renders a true comparison between diseases somewhat difficult. Second, the overall study population is relatively small. Third, items of the medical history may be subjected to recall bias which we mitigated by assessing multiple sources of information (e.g., well-child visits, available smartphone video recordings). Fourth, the actual baseline study assessments may be subject to ascertainment bias, because the diagnosis was known and the investigator was not blinded. Fifth, the assessment of first neurological sign and symptom focused more on the motor system as this is more intuitive to ascertain, therefore more subtle, early non-motor issues may have been missed. Sixth, some items used had limited age ranges (e.g., 9HPT, WHODAS, or SARA scores) and could therefore not be applied in the entire study population. 9HPT turned out not to be suitable in the present population because its use was too complex, whereas the easier SARA assessments were very useful. These limitations are due to the rare nature of the condition. We nevertheless consider our data generalizable within the context of these important limitations. The strength of this study is the systematic protocol-guided assessment by a small team of trained investigators that will be able to capture the longitudinal outcomes of softer endpoints in the future including the variability of changes over time which is relevant for estimations of sample sizes and study durations for future therapeutic trials.

## Conclusion

Data in this registry raise awareness of these rare and fatal conditions in order to accelerate diagnosis, inform counselling of afflicted families, define quantitative endpoints for clinical trials, and could serve as historical controls for future therapeutic studies.

## Supporting information

STROBE checklist

## Data Availability

All data produced in the present study are available upon reasonable request to the author

## Acknowledgment

We thank Lorna Stimson, PhD, for language editing.

## Notes

**Conflict of Interest Notification Page** Dr. Ries declares no conflict of interest. Dr. Mendoza declares no conflict of interest. Dr. Arash-Kaps declares no conflict of interest. Mr. Quack is a volunteering member the patients’ organization “Hand in Hand against Tay-Sachs and Sandhoff disease” who financially supported this study. Mrs. Hardt is a volunteering member the patients’ organization “Hand in Hand against Tay-Sachs and Sandhoff disease” who financially supported this study. Dr. Amraoui declares no conflict of interest. Dr. Beck received honoraria from Takeda. Dr. Mengel received research grants and consultation fees and speakers’ honoraria from Sanofi Genzyme, Amicus, Takeda, Orphazyme, Prevail, Idorsia, Sio and Taysha.

### Competing Interest Statement

Dr. Ries declares no conflict of interest.
Dr. Mendoza declares no conflict of interest.
Dr. Arash-Kaps declares no conflict of interest.
Mr. Quack is a volunteering member the patients' organization "Hand in Hand against Tay-Sachs and Sandhoff disease" who financially supported this study.
Mrs. Hardt is a volunteering member the patients' organization "Hand in Hand against Tay-Sachs and Sandhoff disease" who financially supported this study.
Dr. Amraoui declares no conflict of interest.
Dr. Beck received honoraria from Takeda.
Dr. Mengel received research grants and consultation fees and speakers' honoraria from Sanofi
Genzyme, Amicus, Takeda, Orphazyme, Prevail, Idorsia, Sio and Taysha.

### Funding Statement

This study was jointly funded by the patients' advocacy organization “Hand in Hand e.V.” and the independent non-profit clinical research organization "SphinCS Lyso gemeinnuetzige UG".

### Author Declarations

This study was approved by the state medical board Hessen, Frankfurt, Germany (registration number 2019-1483-evBO) and registered on clinicaltrials.gov (NCT04624789). All parents, caregivers, or patients signed informed consent or, if applicable, assent before inclusion into the study.

