## Supplemental information for "Quantitative longitudinal natural history of eight gangliosidoses – conceptual framework and baseline data of the German 8-in-1 disease registry. A cross-sectional analysis"

Supplemental Table 1

Summary of the conditions by disease group, their respective OMIM phenotype numbers, their underlying biochemical deficiencies, and their incidences. The 8-in-1 registry is intended to capture data on the following eight diseases out of two major groups:

the G_M1_-gangliosidosis / sialidosis group comprises the following conditions: 1) G_M1_-gangliosidosis, 2) Morquio B variant, 3) sialidosis, 4) galactosialidosis.

Likewise, the G_M2_-gangliosidosis group includes: 5) Tay-Sachs disease, 6) Tay-Sachs B1 variant disease, 7) Sandhoff disease, 8) G_M2_-activator deficiency

| Disease group | Disease number | Disease | OMIM Number (Orpha number) | Deficiency | Prevalence (P); Incidence (I); Birth prevalence (BP) ^1^ |
| --- | --- | --- | --- | --- | --- |
| G_M1_- gangliosidosis / sialidosis group | 1 | G_M1_- gangliosidosis | # 230500  (infantile)  # 230600  (juvenile)  #230650  (adult) | Beta-galactosidase | 0,75/ 100.000¹  BP* |
|  | 2 | Morquio B variant | #253010 | Beta-galactosidase | NA |
|  | 3 | Sialidosis | # 256550 | Neuraminidase | 0,05/100.000¹  BP* |
|  | 4 | Galactosialidosis | # 256540 | Protective protein /cathepsin A | NA (100 published cases)¹ |
| G_M2_- gangliosidosis group | 5 | Tay-Sachs disease | # 272800 | Hexosaminidase A | 5/100.000¹ P* |
|  | 6 | Tay-Sachs B1 variant | # 272800 | Hexosaminidase A |  |
|  | 7 | Sandhoff disease | # 268800 | Hexosaminidase A&B |  |
|  | 8 | G_M2_-activator deficiency | # 272750 | G_M2_-activator | NA (10 published cases)¹ |

* An asterisk indicates European data

Supplemental Table 2

8-in-1 score: individual items and attributed point ratings

| Participation | Score |
| --- | --- |
| Unrestricted | 0 |
| Occasional assistance (in kindergarten, school, work) | 1 |
| Constant assistance (in kindergarten, school, work, e.g. by individual helpers or integration force) | 2 |
| Special facility (kindergarten/school/work for intellectually/physically disabled, sheltered workshop) | 3 |
| No participation (no kindergarten, no school, no work/ no sheltered workshop) | 4 |
| Care/Independency | **Score** |
| No impairment of independency, no assistance for hygiene / everyday activities | 0 |
| Slight impairment of independency, little assistance for hygiene / everyday activities | 1 |
| Considerable impairment of independency, often assistance for hygiene / everyday activities | 2 |
| Severe impairment of independency, constant assistance for hygiene / everyday activities, i.a. additional assistance by nursing staff | 3 |
| Most severe impairment of independency, care is carried out entirely by caregiver/ nursing staff (24-hour care) | 4 |
| Ambulation | **Score** |
| Unrestricted | 0 |
| Conspicuous gait ("clumsy", stumbling, falls), no walking assistance | 1 |
| Walking distance ≥ 20 m without assistance | 2 |
| Walking distance < 20 m without assistance | 3 |
| No walking, no steps (wheelchair-dependent, bedridden) | 4 |
| Dexterity/ Fine motor skills | **Score** |
| Unrestricted | 0 |
| Clumsy (self-dependence preserved) | 1 |
| Slight limitations (occasional assistance, e.g. when eating or for other fine motor activities) | 2 |
| Moderate limitations (constant assistance, e.g. when eating or for other fine motor activities) | 3 |
| Severe limitations, functionless (no fine motor skills) | 4 |
| Speech | **Score** |
| Unrestricted | 0 |
| Unclear, but easy to understand | 1 |
| Unclear, difficult to understand | 2 |
| Incomprehensible, vocalization, nonverbal communication | 3 |
| Anarthria, complete loss of speech | 4 |
| Swallowing | **Score** |
| Unrestricted | 0 |
| Conspicuous, slow swallowing or occasional dysphagia | 1 |
| Constant dysphagia with liquids | 2 |
| Constant dysphagia with liquids and solids (i.a. partial enteral nutrition) | 3 |
| Severe dysphagia, complete enteral nutrition | 4 |
| Epilepsy | **Score** |
| No | 0 |
| < 2 seizures per week, controlled with a medication, mostly seizure-free | 1 |
| Frequent seizures (≥ 2 seizures per week, not daily), controlled with 1-2 medications, not seizure-free | 2 |
| Daily seizures, controlled only with 2-3 medications, partial therapy response | 3 |
| Uncontrolled seizures despite drugs, complete resistance to therapy | 4 |
| Cognition | **Score** |
| Unrestricted | 0 |
| Mild intellectual disability (adults: intellectual age 9-12 years, children: learning difficulties at school) | 1 |
| Moderate intellectual disability (adults: intellectual age 6-9 years; children: distinct developmental delay, school for intellectually disabled; adults/ children: reading and writing possible) | 2 |
| Severe intellectual disability (adults: intellectual age 3-6 years; children: distinct developmental delay, special school for intellectually disabled; adults/ children: no reading and writing, incapable of schooling, but educable for practical life, permanent support) | 3 |
| Most severe intellectual disability (adults: intellectual age under 3 years; children: most severe developmental disorder; adults/children: development support no longer possible, incapable of schooling, not educable for practical life, extensive support) | 4 |
| Total score |  |

Supplemental Table 3 (separate document)

Synopsis of the study population and study results.

* - Two patients could not be clearly attributed to a phenotype. According to the classification item "first motor symptom", the two patients would be considered as infantile respectively late infantile ^2^, whereas according the classification item "best motor performance", they would be considered late infantile, and, respectively, juvenile.^2^ We considered one of them late infantile and the other one juvenile according to their best motor performance, because this was most consistent with their observed disease trajectory.^3^

Precise definitions of 8-in-1 subscores are listed in Supplemental Table 2

\# - denotes novel mutation

**- Patients 3 and patients 21-23 were previously mentioned in the published literature.^4,5^

nd: not done

na: not available

Supplemental Figures

Supplemental Figure 1

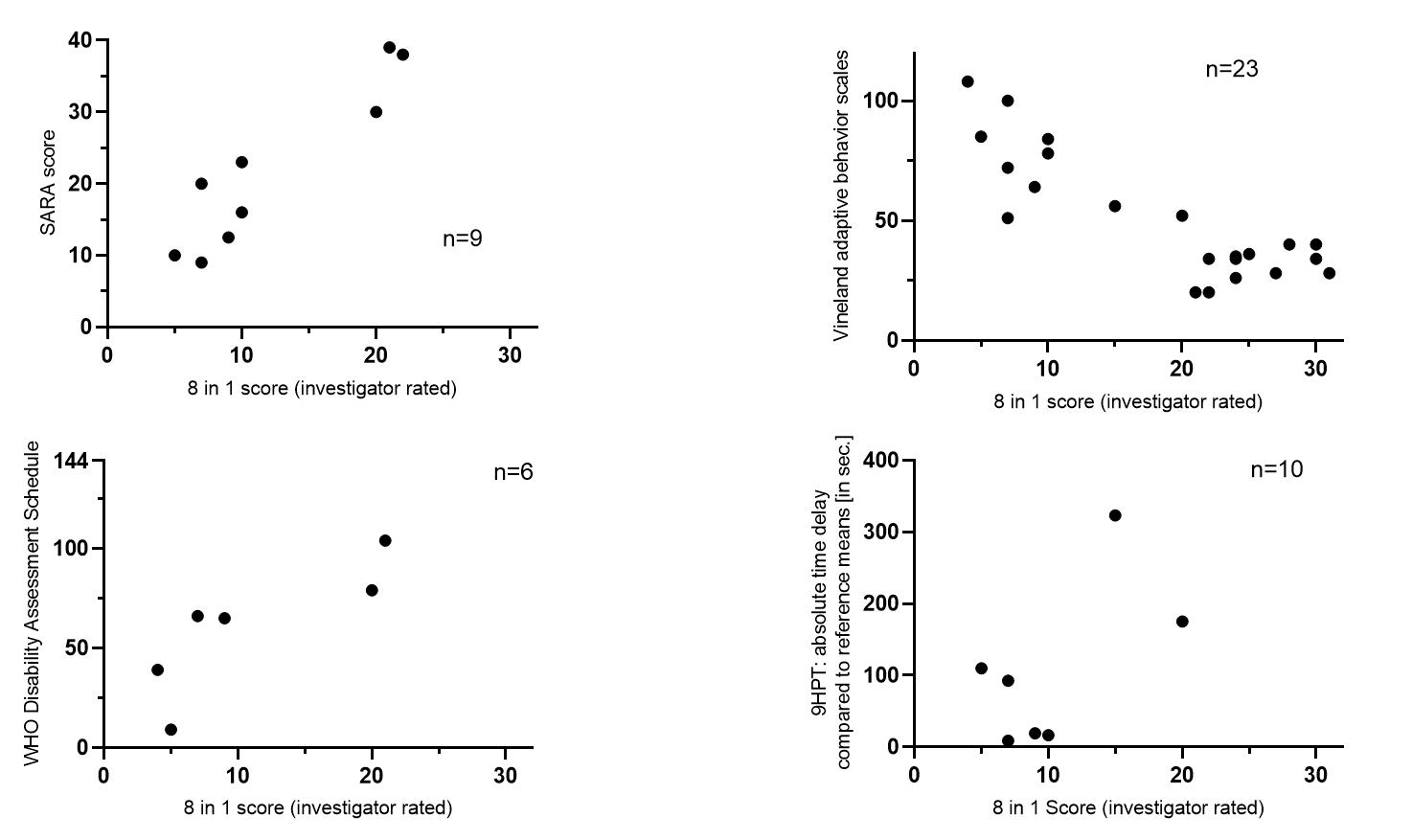

Correlation of 8 in 1 scores with previously validated instruments of a. cognitive development, b. ataxia, c. dexterity, and d overall disability.

Correlation of disease severity (assessed by investigator rated 8 in 1 scores) with validated instruments for **A.** development (Vineland adaptive behavioral scales, Pearsons' r = -0.5312; p=0.0110), **B.** ataxia (SARA scores, Pearsons' r = 0.6806; p=0.0117), **C.** manual dexterity (9 hole peg test, Pearsons' r = 0.4293; p=NS), and **D.** overall disability (WHO disability assessment schedule 2.0, (Pearsons' r = 0.6929; p=NS)

Higher scores indicate more severe disease in 8-in-1 scores, SARA scores, 9 hole peg test performance delay, and WHODAS 2.0 scores. In contrast, higher Vineland scores indicate better overall development in given patient.
